# PROPOSED SURVEY FOR PATIENTS PRIOR TO MAMMOGRAPHY AS A TOOL TO IMPROVE RADIOLOGICAL REPORTING

**DOI:** 10.1101/2024.06.09.24308659

**Authors:** Fernando Muñoz Villaseca, Valeria Pardo Toro, Elisabeth Cañipa Zulueta, Marco Jiménez Herrera, Jeannette Soto Miranda

**Affiliations:** Student, Medical Technology, Central University of Chile; Faculty, Central University of Chile

**Keywords:** breast, mammography, breast neoplasia, risk factor, surveys and questionnaires

## Abstract

**Background:** Breast cancer is one of the leading causes of death among women worldwide. Therefore, early detection through quality mammography, along with information collected from the mammographic survey, is crucial.

The objective of this research is to propose a mammographic survey for patients prior to mammography to gather important information for radiologists specializing in mammography when preparing reports.

**Methodology:** This research was conducted using a semi-structured survey with both closed and open-ended questions, which was completed by 31 radiologists specializing in mammography.

**Results:** Relevant patient history and risk factors were collected, which are most useful for radiological reporting concerning patient history and potential risk factors.

**Conclusion:** Based on the responses collected from radiologists, we propose a final mammographic survey as a tool to aid radiologists in correlating clinical images with patient history. This survey emphasizes family history, gynecological-obstetric history, surgical history, among other factors.

## Background

Breast cancer is an abnormal proliferation of the cells that make up the epithelium of the mammary lobules, which have the capacity to spread. Therefore, breast cancer requires rapid detection and intervention before it disseminates into adjacent tissues. Screening is the only way to detect cancers at an early stage, and thus, it is also capable of reducing deaths from this cancer(Autier & Boniol, 2018; Milosevic et al., 2018), To this end, various techniques are applied, the primary one being mammography (Autier & Boniol, 2018), along with the mammographic survey. Additionally, complementary imaging examinations such as breast ultrasound, breast magnetic resonance imaging (MRI), computed tomography (CT), positron emission tomography (PET), scintigraphy, tomosynthesis, etc., are used.(Le-Petross & Shetty, 2011; Mehnati & Tirtash, 2015).

The mammographic survey is a questionnaire completed prior to the mammogram, serving as the first step in obtaining the patient’s medical history to gather relevant information such as risk factors(Faryabi et al., 2023; Sun et al., 2017). Risk factors include a personal history of benign breast disease, such as fibrocystic mastopathy, non-atypical proliferative lesions, and atypical hyperplasia. These histological precursor lesions have been shown to be associated with an increased risk of subsequent development of breast cancer(Elmore & Gigerenzer, 2005; Hartmann et al., 2005; Sun et al., 2017).

The risk of breast cancer increases with age due to reproductive factors, the use of postmenopausal hormones, weight, and alcohol consumption (Colditz & Rosner, 2000). Early age at menarche and late menopause are also associated with a higher risk of breast cancer (Hsieh et al., 1990).

Another important risk factor is breast density, as it complicates imaging visualization (NF et al., 2007). Additionally, women with dense breasts have a higher risk of breast cancer compared to those with low-density breasts (“Mammographic Breast Density,” 2007; Mandelson et al., 2000; McCormack & Dos Santos Silva, 2006).

Regarding family history, a cohort study of over 113,000 women in the United Kingdom demonstrated that women with a first-degree relative with breast cancer have a 1.75 times greater risk of developing the disease compared to women without affected relatives. Furthermore, the risk is 2.5 times higher or more in women with two or more first-degree relatives with breast cancer (Sun et al., 2017).

The ethnicity has been extensively studied, with the lifetime risks of specific subtypes of breast cancer varying according to race, with luminal breast cancer being more prevalent in white women and triple negative breast cancer in black women (Kurian et al., 2010a). Several studies indicate that ethnic differences correlate with different types of risks (Hughes et al., 1996; Kurian et al., 2010b; *Lifetime Risks of Specific Breast Cancer Subtypes among Women in Four Racial/Ethnic Groups - Consensus*, n.d.; Reeves et al., 2010).

Nulliparity is associated with an increased risk of breast cancer (Clavel-Chapelon & Gerber, 2002), similarly, women who have their first child at an older age, after 35 years old, also face an increased risk (Ewertz et al., 1990). In this line of research, breastfeeding is associated with a decreased risk of breast cancer in women who have breastfed at any point compared to those who have not (Beral et al., 2002).

Hysterectomy with oophorectomy reduces the risk of breast cancer by eliminating the primary source of estrogen. (Rebbeck et al., 2009), Similarly, hysterectomy without oophorectomy also reduces the risk of breast cancer (Bernard et al., 2009).

Other surgeries directly involving the breast, such as partial or total mastectomy, are significant precedents. Local recurrence following mastectomy or breast-conserving surgery significantly impacts the survival of these patients, substantially reducing it (Fodor et al., 2008; Mo et al., 2021; Wu et al., 2021)

Another aspect that can be recorded in a pre-mammography survey is signs and symptoms. Among them is breast self-examination, with the purpose of detecting any breast lump (Abdel-Razeq et al., 2020), nipple or skin retraction, pain, discharge, among others, are also included (Koo et al., 2017; Nyström et al., 1993). Important points to consider for recording in the patient’s medical history, as some of these characteristics may be associated with malignancy.

Given this background, the purpose of this study is to gather the opinions of radiologists specializing in mammography through a questionnaire and propose a pre-mammography survey incorporating the most important risk factors gathered from the literature and the expertise of specialists. The aim is to provide radiologists with more information when generating radiological reports, thereby enhancing the early detection of breast cancer.

## Materials and Methods

This study has been approved by the ethics committee of our institution. In this descriptive, observational study, questions related to the risk factors associated with breast cancer, as described in the literature, and outlined in the introduction, were formulated.

Survey: Semi-structured with closed and open-ended questions, where responses are measured nominally. Closed-ended questions contain predetermined response categories or options, presenting participants with limited choices to select from. On the other hand, open-ended questions do not predefine response alternatives, allowing for a wide range of possible answers. In this case, the main part of the survey consists of dichotomous closed-ended questions, with only two possible responses: “yes” and “no.” Following each closed-ended question, participants were given the option to answer an open-ended question to provide additional information and elaborate on their opinions.

The responses collected from the surveys were coded using the following procedure:

1. Select the number of responses through an appropriate sampling method to ensure the representativeness of the surveyed participants.
2. Observe the frequency of each response to the questions.
3. Choose the responses that appear most frequently.
4. Classify the responses according to a logical criterion, ensuring they are mutually exclusive.
5. Assign a title to each theme that is representative.
6. Assign a code to each general response pattern.

### The questions were as follows

Question #1: Regarding the patient’s family history, which one or ones do you consider useful for the radiological report? Mark YES/NO.

Question #2: Regarding the patient’s gynecological and obstetric history, which one or ones do you consider useful for the radiological report? Mark YES/NO.

Question #3: Regarding the patient’s surgical history, which one or ones do you consider useful for the radiological report? Mark YES/NO.

Question #4: Regarding the patient’s signs and symptoms, which one or ones do you consider useful for the radiological report? Mark YES/NO.

Question #5: Regarding the patient’s modifiable risk factors, which one or ones do you consider useful for the radiological report? Mark YES/NO.

Question #6: Do you consider it useful for the radiological report to know the patient’s ethnicity? Mark YES/NO.

Question #7: Do you consider the following images representative for the mammography report? Mark YES/NO.

Additionally, 5 open-ended questions were asked following questions #1, #2, #3, #4, and #5 respectively, giving radiologists the opportunity to add information about unmentioned background factors they consider useful for the radiological report. It’s worth noting that in these questions, respondents could mention more than one background factor.

Question #7 presented 4 options of representative breast images. Image 1 (Figure 1) displays the anteroposterior breasts, showing the mammary quadrants, axillary area, and neck.

**Figure 1:**
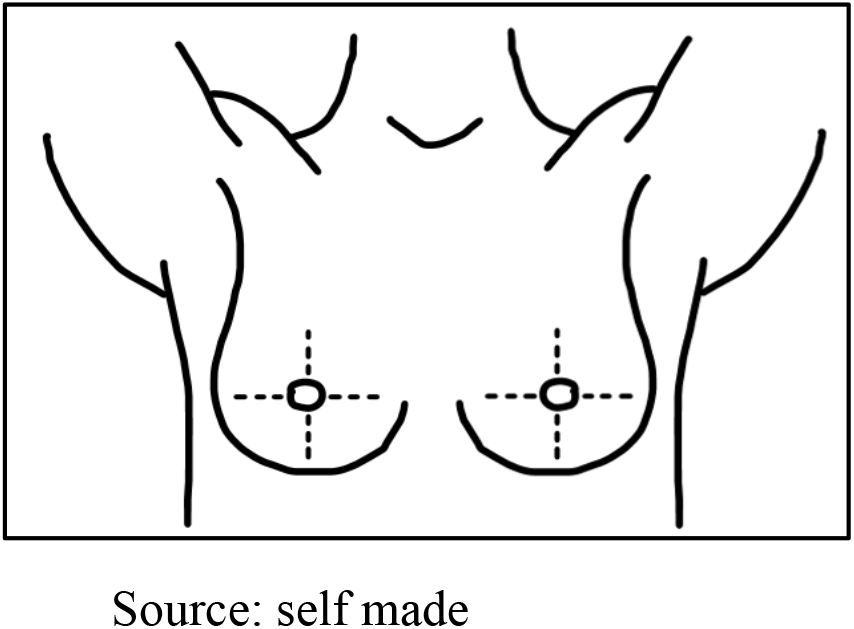
Image 1

Image 2 (Figure 2) depicts the anteroposterior view of the breasts, displaying the mammary quadrants, and the lateral view showing the mammary thirds.

**Figure 2:**
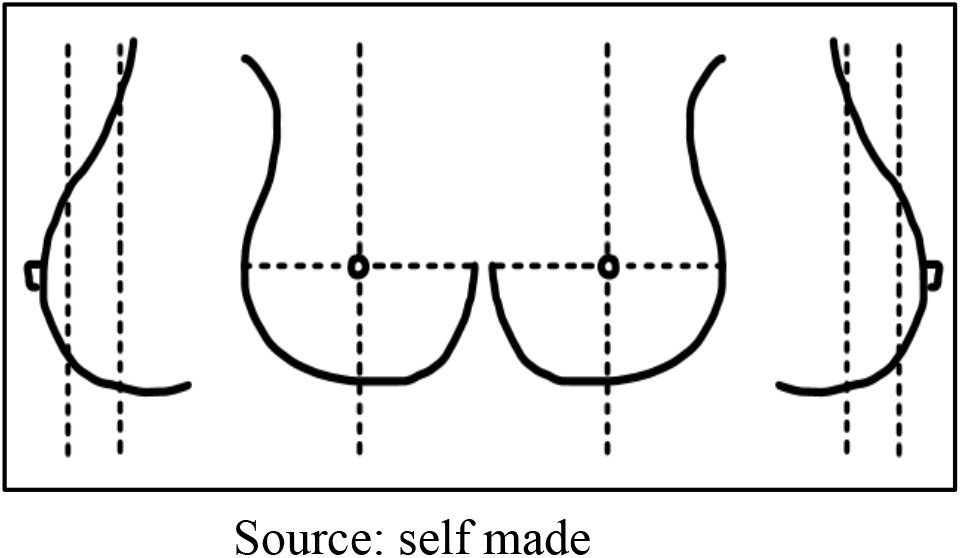
Image 2

Image 3 (Figure 3) displays both the anteroposterior and lateral views of the breasts, with the anteroposterior view showing the mammary quadrants.

**Figure 3:**
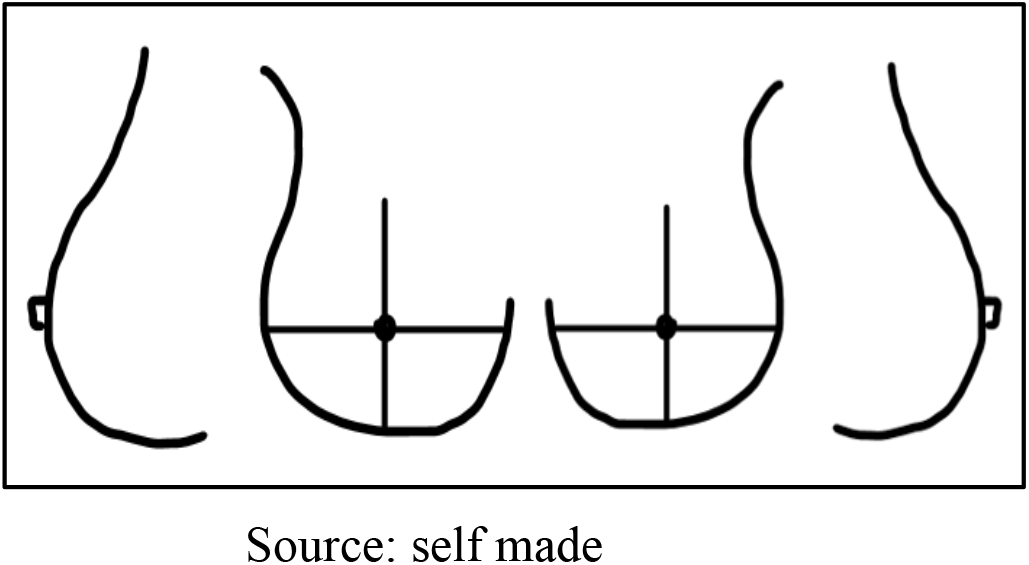
Image 3

Image 4 (Figure 4) only shows the breasts in anteroposterior and lateral views, without the quadrants and mammary thirds.

**Figure 4:**
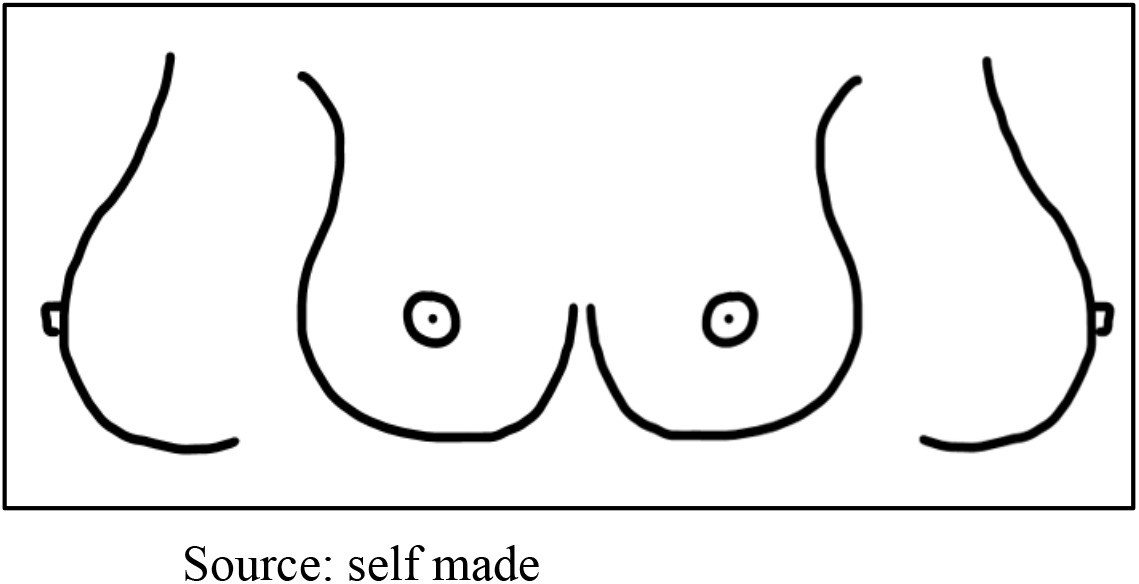
Image 4

Subsequently, it was distributed via the Google Forms platform to 38 radiologists across Chile, resulting in 31 responses out of the 38 surveys sent. Informed consent was obtained online from each participant prior to the survey, conducted through Google Forms, ensuring confidentiality of information, data identification, and voluntary participation.

Data reporting primarily utilized frequencies and percentages to identify the most important risk factors according to the respondents.

## Results

### Family History of the Patient

Question #1: Regarding the patient’s family history, which ones do you consider useful for the radiological report?

- 100% (31 respondents) consider it useful to know if the patient’s mother and sister have a history of breast cancer.
- 93.5% (29 respondents) consider it useful to know if the patient’s grandmother and daughter have a history of breast cancer.
- 80.6% (25 respondents) consider it useful to know if the patient’s aunt has a history of breast cancer.
- 54.8% (17 respondents) consider it useful to know if the patient’s cousin has a history of breast cancer.

Regarding question #1, do you consider any other unmentioned family history?

24 respondents considered an unmentioned family history in question #1, as follows:

- 10 respondents considered paternal lineage of breast cancer.
- 5 respondents considered a family history of ovarian cancer.
- 4 respondents considered the age of the family member with breast cancer.
- 4 respondents considered male breast cancer history.
- 1 respondent considered family history of genetic mutation.

### Gynecological and obstetric background of the patient

Question #2: Regarding the patient’s gynecological and obstetric history, which ones do you consider useful for the radiological report?

- 100% (31 respondents) consider it useful to know if the patient has undergone hormone replacement therapy and if she has taken tamoxifen.
- 96.7% (30 respondents) consider it useful to know if the patient breastfed (lactation).
- 93.5% (29 respondents) consider it useful to know if the patient is currently pregnant.
- 83.8% (26 respondents) consider it useful to know if the patient’s menopause was natural.
- 80.6% (25 respondents) consider it useful to know if the patient has undergone ovariectomy.
- 74.1% (23 respondents) consider it useful to know the age of menarche and if the patient uses a contraceptive method.
- 67.7% (21 respondents) consider it useful to know if the patient’s menopause was surgical.
- 64.5% (20 respondents) consider it useful to know the age of the patient’s first pregnancy.
- 54.8% (17 respondents) consider it useful to know the number of children the patient has.
- 48.3% (15 respondents) consider it useful to know the patient’s last menstrual period (FUR).
- 25.8% (8 respondents) consider it useful to know if the patient has undergone a hysterectomy.
- 22.5% (7 respondents) consider it useful to know the number of deliveries the patient has had.

Regarding question #2, do you consider any other unmentioned history?

Five respondents mentioned another gynecological-obstetric history not mentioned in question #2. These are:

- 2 respondents considered the age of menopause.
- 2 respondents considered ovarian cancer.
- 1 respondent considered post-menopausal obesity.

Question #3, which surgical history or histories do you consider useful for the radiological report?

- 100% (31 respondents) consider it useful to know if the patient has undergone partial mastectomy, total mastectomy, stereotactic biopsy, excisional biopsy, and radiotherapy.
- 96.7% (30 respondents) consider it useful to know if the patient has undergone core biopsy, breast augmentation or reduction surgery, and chemotherapy.
- 93.5% (29 respondents) consider it useful to know the composition of the breast implant (silicone-saline).
- 67.7% (21 respondents) consider it useful to know if the patient has had mastitis.
- 61.2% (19 respondents) consider it useful to know if the patient has undergone aspiration biopsy.
- 54.8% (17 respondents) consider it useful to know the position of the breast implant.

Regarding question #3, which surgical history or histories do you consider useful for the radiological report?

- 100% (31 respondents) consider it useful to know if the patient has undergone partial mastectomy, total mastectomy, stereotactic biopsy, excisional biopsy, and radiotherapy.
- 96.7% (30 respondents) consider it useful to know if the patient has undergone core biopsy, breast augmentation or reduction surgery, and chemotherapy.
- 93.5% (29 respondents) consider it useful to know the composition of the breast implant (silicone-saline).
- 67.7% (21 respondents) consider it useful to know if the patient has had mastitis.
- 61.2% (19 respondents) consider it useful to know if the patient has undergone aspiration biopsy.
- 54.8% (17 respondents) consider it useful to know the position of the breast implant.

Regarding question #3, do you consider any other unmentioned surgical history?

Fourteen respondents considered another surgical history not mentioned in question #3. These are:

- 4 respondents considered biopsy results.
- 3 respondents considered surgical history of any other cancer.
- 2 respondents considered lipotransfer.
- 2 respondents considered axillary surgery.
- 1 respondent considered neoadjuvant clip.
- 1 respondent considered dates of submitted surgeries.

### Signs and symptoms

Question #4, here are the findings regarding signs and symptoms considered useful for the radiological report:

- 100% (31 respondents) consider it useful to know if the patient presents tumor formation, skin retraction, and discharge.
- 96.7% (30 respondents) consider it useful to know if the patient presents skin thickening and inverted nipple.
- 83.8% (26 respondents) consider it useful to know if the patient presents no symptoms (asymptomatic).
- 70.9% (22 respondents) consider it useful to know if the patient presents breast pain.

Regarding question #4, 12 respondents considered additional signs and symptoms not mentioned in the question. These are:

- 4 respondents considered erythema.
- 4 respondents considered axillary anomalies.
- 2 respondents considered adenopathy.
- 2 respondents considered the duration of symptoms.
- 1 respondent considered eczema.
- 1 respondent considered edema.
- 1 respondent considered ulceration.
- 1 respondent considered fistula.
- 1 respondent considered moles.
- 1 respondent considered breast malformation.

Modifiable Risk Factors of the Patient

Question #5: Regarding the modifiable risk factors of the patient, which one(s) do you consider useful for the radiological report?

- 74.1% (23 respondents) consider it useful for the radiological report to know if the patient is obese.
- 64.5% (20 respondents) consider it useful for the radiological report to know if the patient smokes tobacco.
- 61.2% (19 respondents) consider it useful for the radiological report to know if the patient consumes alcohol.
- 48.3% (15 respondents) consider it useful for the radiological report to know if the patient leads a sedentary lifestyle.
- 32.2% (10 respondents) consider it useful for the radiological report to know the patient’s diet.

Regarding question #5, do you consider any other unmentioned factors? 2 respondents indicated that these factors are not relevant for the radiological report but are relevant for the patient’s clinical assessment.

### Ethnicity of the patient

Question #6: Do you consider it useful for the radiological report to know the ethnicity of the patient?

54.8% (17 respondents) consider it useful for the radiological report to know the ethnicity of the patient.

### Representative images for mammography report

Question #7: Do you consider the following images representative for the radiological report? (see images 1, 2, 3, and 4)

90.3% (28 respondents) consider image 1 useful for the radiological report. 48.3% (15 respondents) consider image 2 useful for the radiological report. 22.5% (7 respondents) consider image 3 useful for the radiological report. 12.9% (4 respondents) consider image 4 useful for the radiological report.

## Conclusions and Discussion

Regarding family history, mammography specialist radiologists consider that the most useful risk factors are knowing if the patient has a family history of breast cancer from the mother (100%), sister (100%), grandmother (93.5%), daughter (93.5%), maternal and/or paternal aunt (80.6%), and history of ovarian cancer. Knowing the age at the time of cancer detection.

This is one of the most important factors to consider and coincides with (Sun et al., 2017), which indicates that the probability increases 1.75 times with one family member and 2.5 times if there are two or more relatives with a history.

Regarding gynecological and obstetric history, mammography specialist radiologists consider that the most useful risk factors are hormone replacement therapy (HRT) (100%), use of tamoxifen (100%), breastfeeding (96.7%), natural menopause (83.8%), ovariectomy (80.6%), age at menarche (74.1%), contraceptive use (74.1%), surgical menopause (67.7%), age at first pregnancy (64.5%), and number of children (54.8%), knowing the dates of each history. The number of children, especially nulliparity, increases the risk of breast cancer according to (Clavel-Chapelon & Gerber, 2002), similarly, hormone intake as studied by (Colditz & Rosner, 2000). Breastfeeding is a protective factor according to (Beral et al., 2002), that is how (Rebbeck et al., 2009) considered that hysterectomy and oophorectomy reduce the risk of breast cancer since they are the main precursors of estrogen in women, which must be considered in the mammographic survey. Regarding the patient’s surgical history, mammography specialist radiologists consider that the most useful risk factors to know are if the patient has undergone partial mastectomy (100%), total mastectomy (100%), stereotactic biopsy (100%), excisional biopsy (100%), radiotherapy (100%), core biopsy (96.7%), breast augmentation or reduction surgery (96.7%), chemotherapy (96.7%), mastitis (67.7%), fine needle aspiration (61.2%), and implant position (54.8%), knowing the dates of each event. All these antecedents are of utmost importance to the respondents. It is well known that recurrence after partial or total mastectomy reduces survival rates, as indicated (Fodor et al., 2008), which is an important concept to know prior to mammography.

Regarding the signs and symptoms of the patient, mammography specialist radiologists consider the most useful ones to know are if the patient is asymptomatic (83.8%) or if she presents with a lump (100%), skin retraction (100%), discharge (100%), thickening of the skin (96.7%), inverted nipple (96.7%), and pain (70.9%), also knowing the dates of onset for each sign and symptom. These signs and symptoms are direct evidence of a possible risk of breast cancer, although there may be benign or malignant findings, they should be taken into consideration when making the diagnosis with mammography, as these antecedents are well identified in the literature regarding the presence of a lump (Abdel-Razeq et al., 2020), and skin retraction, pain, or discharge (Koo et al., 2017; Nyström et al., 1993).

The radiologist specialists in mammography consider that the most useful modifiable risk factors for the patient are obesity (74.1%), smoking (64.5%), and alcohol consumption (61.2%), which aligns with studies evaluating these risks and reporting consistent results, affirming that these factors increase the risk of breast cancer (Colditz & Rosner, 2000).

Regarding the ethnicity factor, radiologists consider it useful to know the patient’s ethnicity (54.8%). There is ample literature indicating that depending on ethnicity, there are differences in the risk of breast cancer. (Hughes et al., 1996; Kurian et al., 2010a, 2010b; Reeves et al., 2010). This makes it necessary to know the patient’s ethnicity beforehand.

Regarding representative images for the radiological report, radiologists consider image 1 (90.3%) to be the most useful.

(The final survey model is available in supplementary material).

The disadvantages of this study include the uncertainty regarding the reach of this survey among radiologists throughout our country, as we lack data on the number of specialist mammography radiologists and how to contact them. The formulated questions may not be sufficient, but this survey can expand in future proposals.

The advantages of this work lie in successfully gathering information from a large number of specialist doctors in this field, unlike any other study, which lends it significant validity. Another advantage is the evidence found, providing ample information to create the final proposal. Furthermore, proposing a survey usable in any imaging center is advantageous, as the included risk factors align with literature from various parts of the world. Having prior knowledge of these factors is crucial for a clear understanding of potential findings in the image and enables the radiologist to make a more informed diagnosis with concrete data.

## Data Availability

All data produced in the present study are available upon reasonable request to the authors

